# Overweight and hypertension profiles among young smokers: a secondary data analysis of the Indonesian National Health Survey 2023

**DOI:** 10.64898/2026.02.06.26345724

**Authors:** Rohana Uly Pradita Siregar, Yoerdy Agusmal Saputra, Vether Fernhandho, Amelia Dyah Kartika Sari

**Affiliations:** Universitas Pelita Harapan; Universitas Sriwijaya; Ministry of Population and Family Development of the Republic of Indonesia

## Abstract

**Background:** Tobacco use among young people remains a major public health concern in Indonesia, where smoking prevalence is high and metabolic risk factors such as overweight and hypertension are increasing. Evidence linking smoking behavior, particularly e-cigarette use, to early cardiometabolic risk in low- and middle-income countries is still limited. This study examined overweight and hypertension profiles among young smokers using nationally representative data from Indonesia.

**Methods:** This secondary analysis used data from the Indonesian Health Survey 2023. Participants were young adults aged 18–25 years with complete information on smoking status, anthropometry, and blood pressure (n = 12,770). Smoking status was categorized as conventional smokers, e-smokers, and dual smokers. Outcomes included overweight/obesity (BMI ≥23 kg/m^2^), central obesity (waist circumference ≥90 cm for men and ≥80 cm for women), and hypertension (≥130/80 mmHg). Logistic regression models estimated adjusted odds ratios (AOR) controlling for age, gender, smoking duration, residence, and socioeconomic proxy variables.

**Results:** Most respondents were conventional smokers (94%), followed by dual smokers (4%) and e-smokers (2%). E-smokers showed higher mean BMI and the greatest prevalence of overweight/obesity (40%) and central obesity (18%). After adjustment, e-smokers and dual smokers had higher odds of overweight/obesity (AOR = 1.37 and 1.41, respectively) and central obesity (AOR = 1.47 and 1.53, respectively) compared with conventional smokers. Hypertension prevalence (11–13%) did not differ significantly across smoking categories.

**Conclusion:** Among young Indonesian smokers, e-cigarette and dual use were associated with higher odds of overweight and central obesity but not hypertension. These findings highlight the importance of integrating tobacco control with early metabolic risk prevention strategies targeting youth.

## Introduction

Tobacco use remains a leading global health threat, causing on the order of 7–8 million deaths each year.^1,2^ In the 2019 Global Burden of Disease analysis, smoking was responsible for about 7.7 million deaths and was the first risk factor for male mortality worldwide.^3^ The adverse effects of tobacco on cardiovascular and metabolic health are well established.^4–6^ Smoking promotes atherosclerosis and is a major cause of heart attack, stroke, and peripheral vascular disease.^3,5^ It also fosters insulin resistance and type 2 diabetes—indeed, tobacco smoking was identified as the third largest global risk factor for diabetes burden.^7^ In aggregate, smokers have higher rates of metabolic syndrome, hypertension, dyslipidemia, and glucose intolerance compared to non-smokers.^6–8^ Mechanistically, nicotine acutely suppresses appetite and raises basal metabolism,^9,10^ which often leads smokers to have lower body weight or BMI than non-smokers.^10^ Paradoxically, however, smoking shifts fat distribution unfavorably: smokers tend to accumulate visceral (abdominal) fat despite lower overall weight.^9,11^ Experimental studies show nicotine increases adipose tissue lipolysis and frees fatty acids, inducing insulin resistance,^9,12,13^ while simultaneously stimulating sympathetic tone and thermogenesis.^14^ The net result is that long-term smokers often develop an “unhealthy lean” phenotype—lower BMI but greater central adiposity and higher blood pressure—which increases cardiovascular risk.^9,14,15^

Adolescence and early adulthood are critical periods for habit formation, and smoking initiated at a young age carries long-lasting consequences.^16–18^ Approximately 20% of people remain addicted to tobacco.^19^ Although the number of adult smokers globally has declined (from around 1.38 billion in 2000 to 1.2 billion in 2024), the industry’s strategy of shifting to alternative products (vapes, nicotine pouches) risks creating a new wave of addiction among adolescents.^19,20^ This trend varies greatly between countries and regions: some countries show a marked decline in youth smoking; others—including certain high-income countries—still report relatively high prevalence among youth or an increase in the use of e-products.^21^ Emerging international data suggest that even among youth, smokers begin to show early signs of metabolic dysfunction.^8^ Studies in adolescents have linked tobacco use to components of the metabolic syndrome. For example, reviews note that smoking adversely affects abdominal obesity and blood pressure in adolescents and young adults.^6^ Likewise, young smokers often display the same paradoxical body composition pattern seen in adults—reduced subcutaneous fat but increased central (visceral) fat deposition^9,14^—which can raise insulin resistance and cardiovascular risk even in the absence of frank obesity. In sum, international research (mostly from high-income countries) indicates that youth smoking can be associated with higher waist circumference and elevated blood pressure compared to non-smokers.^6,9^ However, such studies are limited and sometimes contradictory, and they rarely examine long-term outcomes in low- and middle-income settings. Notably, a recent multi-country survey in LMICs found that adolescent tobacco use was not consistently linked to higher BMI, underscoring that the relationship may differ by context.^22^ Overall, there is a paucity of evidence specifically from LMICs on how youth smoking relates to central obesity or hypertension.

Indonesia has one of the world’s highest youth smoking rates. The 2019 Indonesian Global Youth Tobacco Survey (GYTS) shows that the prevalence of active smokers among male adolescents is 38.3%, the proportion who have ever smoked is 67.0%, and the proportion of smokers with high daily consumption (≥2 cigarettes/day) is 39.1%.^23^ On average, smoking begins in the mid- to late teens (around age 16–18), with many youths using clove-flavored (“*kretek*”) or hand-rolled cigarettes due to their low cost and wide availability. E-cigarette use is also rising: a recent survey found about 13.3% of Indonesian high-school–aged youth (15–24 years) were current vapers, and national data show e-cigarette prevalence in 10–18-year-olds climbing from 0.06% (2018) to 0.13% (2023).^24^ Meanwhile, the burden of overweight, obesity, and hypertension is growing among young Indonesians. For example, the 2018 Basic Health Research found that among age group 20–24 years old, about 8% were overweight and 12% were obese.^25,26^ National analyses also report that roughly 4.1% of young adults (ages 26–35) have hypertension, with overweight (BMI ≥ 25) strongly predicting higher blood pressure.^27^ Taken together, these trends—high smoking prevalence in teens and rising metabolic risk in youth—highlight a potentially important public health issue. Investigating how tobacco use intersects with adiposity and blood pressure in Indonesian youth could inform strategies to curb the early development of chronic disease.

Despite these concerns, research in Indonesia has not yet linked youth smoking with metabolic health outcomes. Most Indonesian tobacco studies focus on adult prevalence or on determinants of youth smoking behavior (peers, advertising, etc.),^28^ rather than on physiological effects. No published studies have concurrently examined tobacco use and measures of adiposity or blood pressure in Indonesian adolescents or young adults. Data on emerging products are also sparse: apart from usage surveys,^24^ there is little information on how e-cigarettes or roll-your-own cigarettes relate to obesity or hypertension in this age group. More broadly, global reviews have noted that the bodyweight implications of adolescent smoking remain poorly understood, especially in LMIC contexts.^22^ In sum, an integrated analysis of smoking, overweight/obesity, and blood pressure in Indonesian youth—using recent national survey data—is clearly needed to fill this gap and guide prevention efforts.

The objectives of this study are centered on understanding the association between smoking behavior and indicators of overweight and hypertension among young adults in Indonesia. Specifically, this research aims to examine the profiles of overweight and hypertension among young smokers, providing insights into how tobacco use may be associated with key health risks in this demographic group. By identifying patterns in body weight and blood pressure among smokers, the study seeks to contribute to early prevention efforts and inform public health strategies targeting youth health behaviors.

This analysis will utilize nationally representative data from the National Health Survey, ensuring that the findings accurately reflect the health status of Indonesia’s youth population. The results are expected to provide crucial evidence for policymakers and public health practitioners in designing early prevention programs targeting obesity and hypertension among young people. Ultimately, this study aims to inform the development of integrated public health interventions that combine tobacco control initiatives with broader NCD prevention strategies, supporting Indonesia’s purpose to build a healthier and more resilient young generation.

## Methods

This study utilized data from the Survei Kesehatan Indonesia (SKI) 2023, a nationally representative health survey conducted by the Ministry of Health of Indonesia. SKI 2023 employs a multistage stratified sampling design to capture demographic, behavioral, and clinical health indicators of the Indonesian population. For the present analysis, adults aged 18-25 years with complete information on smoking status, anthropometric measurements, blood pressure, and covariates were included. Individuals who were pregnant or had missing values across key variables were excluded.

Smoking status was categorized into three groups based on self-reported behavior: conventional smokers (those who reported smoking combustible cigarettes only), e-smokers (those who reported exclusive use of electronic cigarettes), and dual smokers (users of both combustible and electronic cigarettes).

Anthropometric outcomes were derived from standardized measurements collected by trained survey officers. Overweight/obesity was defined using the WHO Asian cut-off for BMI, where BMI ≥23 kg/m^2^ indicated overweight or obesity. Central obesity was assessed using waist circumference, with cut-off values of ≥90 cm for men and ≥80 cm for women, consistent with regional recommendations for Asian populations. Blood pressure was measured using calibrated digital sphygmomanometers following a standardized resting protocol. Hypertension was defined as systolic blood pressure ≥130 mmHg, and/or diastolic blood pressure ≥80 mmHg.

Covariates included age (continuous), gender, duration of smoking (years), urban/rural residence, and receipt of government financial aid as a proxy for socioeconomic status. These variables were selected a priori based on theoretical relevance to both smoking behavior and cardiometabolic risk.

All analyses were conducted using SPSS version 30.0. Associations between smoking category and each health outcome (overweight/obesity, central obesity, and hypertension) were examined using logistic regression. Unadjusted odds ratios (OR) were first estimated, followed by adjusted odds ratios (AOR) controlling for the covariates listed above. The conventional smoker group was treated as the reference. Statistical significance was set at p < 0.05, and all analyses accounted for the complex survey design of SKI 2023 through appropriate weighting procedures.

The SKI 2023 survey protocol received ethical approval from the Indonesian Ministry of Health. Written informed consent was obtained from all adult participants. The current study utilized anonymized secondary data provided by the Ministry of Health, therefore, no additional ethical approval was required.

## Results

A total of 12,770 young smokers aged 18–25 years in Indonesia were analyzed, the majority of whom were conventional smokers (about 94%), with smaller proportions of e-smokers (2%) and dual smokers (4%). The characteristics of respondents across the three smoking categories are summarized in Table 1. Although age differences across groups were modest, dual smokers were slightly younger on average. E-smokers differed most from the other groups sociodemographically. For instance, four out of five lived in urban areas, compared with just over half of conventional smokers. They also had a higher proportion of female users (nearly 4%) relative to the predominantly male profile seen among other smoking types. E-smokers tended to start smoking later (around 17.6 years) and had a shorter smoking duration than conventional and dual smokers, indicating a generally more recent adoption of smoking behavior.

**Table 1.** Characteristics of young smokers.

|  | Conventional Smoker | E-Smoker | Dual Smoker | Total | p-value |
| --- | --- | --- | --- | --- | --- |
| N | 11995 | 240 | 535 | 12770 |  |
| Age (years) | 21.76 ± 2.25 | 21.69 ± 2.08 | 21.45 ± 2.22 | 21.75 ± 2.25 | 0.007 |
| Age at Smoking Initiation (years) | 16.02 ± 2.64 | 17.62 ± 2.87 | 15.49 ± 3.01 | 16.02 ± 2.68 | <0.001 |
| Duration of Smoking (years) | 4.37 ± 2.92 | 3.03 ± 2.44 | 4.10 ± 2.93 | 4.33 ± 2.92 | <0.001 |
| Gender |  |  |  |  |  |
| Female | 109 (0.9) | 9 (3.8) | 5 (0.9) | 123 (1.0) | <0.001 |
| Male | 11886 (99.1) | 231 (96.3) | 530 (99.1) | 12647 (99.0) |  |
| Occupation Status |  |  |  |  |  |
| Student | 1693 (14.1) | 54 (22.5) | 92 (17.2) | 1839 (14.4) | <0.001 |
| Employed | 7166 (59.7) | 141 (58.8) | 326 (60.9) | 7633 (59.8) |  |
| Unemployed/Not in school | 3136 (26.1) | 45 (18.8) | 117 (21.9) | 3298 (25.8) |  |
| Area |  |  |  |  |  |
| Urban | 6473 (54.0) | 193 (80.4) | 385 (72.0) | 7051 (55.2) | <0.001 |
| Rural | 5522 (46.0) | 47 (19.6) | 150 (28.0) | 5719 (44.8) |  |
| Receiving Financial Aid |  |  |  |  |  |
| Yes | 3467 (28.9) | 37 (15.4) | 127 (23.7) | 3631 (28.4) | <0.001 |
| No | 8528 (71.1) | 203 (84.6) | 408 (76.3) | 9139 (71.6) |  |
| Comorbid Conditions |  |  |  |  |  |
| Cancer | 1 (0.0) | 0 (0.0) | 0 (0.0) | 1 (0.0) | 0.939 |
| Diabetes | 1 (0.0) | 0 (0.0) | 1 (0.0) | 2 (0.0) | 0.157 |
| Heart Disease | 3 (0.0) | 0 (0.0) | 0 (0.0) | 3 (0.0) | 0.829 |
| Stroke | 1 (0.0) | 0 (0.0) | 0 (0.0) | 1 (0.0) | 0.939 |
| Chronic Kidney Disease | 1 (0.0) | 0 (0.0) | 0 (0.0) | 1 (0.0) | 0.939 |

As shown in Table 2, distinct anthropometric patterns emerged across smoking categories. E-smokers had a noticeably higher mean BMI (22.9 kg/m^2^) than conventional and dual smokers, and they also exhibited the highest prevalence of overweight or obesity, affecting 40% of their group. Dual smokers showed an intermediate pattern, whereas conventional smokers had the lowest proportion with excess weight. A similar trend was evident for central obesity, nearly 18% of e-smokers met the criteria, compared with about 10% of conventional smokers. These findings indicate that adiposity-related markers were more concentrated among e-cigarette users, whether exclusive or dual.

**Table 2.** Distribution of overweight/obesity and hypertension among young smokers.

|  |  |  | Conventional Smoker | E-smoker | Dual Smoker | Total | p-value |
| --- | --- | --- | --- | --- | --- | --- | --- |
| Body Mass Index (kg/m<sup>2</sup>) |  |  | 21.64 ± 3.87 | 22.87 ± 4.67 | 21.91 ± 4.41 | 21.68 ± 3.92 | <0.001 |
| Underweight |  |  | 2301 (19.2) | 33 (13.8) | 124 (23.2) | 2458 (19.2) | <0.001 |
| Normal |  |  | 6157 (51.3) | 111 (46.3) | 227 (42.4) | 6495 (50.9) |  |
| Overweight/Obese |  |  | 3527 (29.5) | 96 (40.0) | 184 (34.4) | 3817 (29.9) |  |
| Waist Circumference (cm) |  |  | 76.21 ± 10.25 | 80.36 ± 12.67 | 76.86 ± 11.70 | 76.31 ± 10.38 | <0.001 |
| Normal |  |  | 10797 (90.0) | 197 (82.1) | 457 (85.4) | 11451 (89.7) | <0.001 |
| Central Obesity |  |  | 1198 (10.0) | 43 (17.9) | 78 (14.6) | 1319 (10.3) |  |
| Systolic Blood Pressure (mmHg) |  |  | 118.41 ± 10.99 | 118.12 ± 11.20 | 118.75 ± 11.65 | 118.42 ± 11.02 | 0.714 |
| Diastolic Blood Pressure (mmHg) |  |  | 78.44 ± 8.59 | 78.98 ± 9.18 | 77.76 ± 9.00 | 78.42 ± 8.62 | 0.121 |
| Normal |  |  | 4633 (38.6) | 97 (40.4) | 227 (42.4) | 4957 (38.8) | 0.192 |
| Pre-hypertension |  |  | 6034 (50.3) | 111 (46.3) | 245 (45.8) | 6390 (50.0) |  |
| Hypertension |  |  | 1328 (11.1) | 32 (13.3) | 63 (11.8) | 1423 (11.1) |  |

Blood pressure profiles, however, showed little variation across groups. Mean systolic pressure was similar for all smokers, around 118 mmHg, and the distribution of hypertension did not differ significantly. Hypertension affected roughly 11–13% of each group, with e-smokers and dual smokers only slightly higher than conventional smokers. Pre-hypertension accounted for about half of respondents regardless of smoking category. This contrasts with the clearer anthropometric differences and suggests that weight-related distinctions were more prominent than blood pressure differences in this age range.

When overweight/obesity and hypertension were evaluated together (Table 3), the proportion affected remained small, only about 2% overall, but was highest among dual smokers (nearly 4%) and lowest among conventional smokers (2%). Most respondents (over 80%) had normal BMI, waist circumference, and blood pressure simultaneously, although e-smokers had a smaller proportion within these normal ranges.

**Table 3.** Combined distribution of overweight/obesity and hypertension among young smokers.

|  |  |  | Conventional Smoker | E-smoker | Dual Smoker | Total | p-value |
| --- | --- | --- | --- | --- | --- | --- | --- |
| Overweight/Obese & Hypertension |  |  | 241 (2.0) | 8 (3.3) | 21 (3.9) | 270 (2.1) | <0.001 |
| Overweight/Obese |  |  | 792 (6.6) | 30 (12.5) | 52 (9.7) | 874 (6.8) |  |
| Hypertension |  |  | 1087 (9.1) | 24 (10.0) | 42 (7.9) | 1153 (9.0) |  |
| Normal BMI, Waist Circumference, and Blood Pressure |  |  | 9875 (82.3) | 178 (74.2) | 420 (78.5) | 10473 (82.0) |  |

Multivariable analysis in Table 4 supported the unadjusted patterns. After controlling for age, gender, smoking duration, area, and financial aid, e-smokers had approximately 1.37 times higher odds of being overweight or obese compared with conventional smokers, while dual smokers had 1.41 times higher odds. A similar pattern was observed for central obesity, where adjusted odds were 1.47 times higher for e-smokers and 1.53 times higher for dual smokers. In contrast, neither e-smokers nor dual smokers demonstrated increased odds of hypertension after adjustment, reflecting the minimal group differences observed in blood pressure indicators.

**Table 4.** Multivariate analysis of overweight/obesity and hypertension.

|  | Overweight/Obese |  | Central Obesity |  | Hypertension |  |
| --- | --- | --- | --- | --- | --- | --- |
|  | OR | AOR<sup>a</sup> | OR | AOR<sup>a</sup> | OR | AOR<sup>a</sup> |
| Conventional Smoker | ref | ref | ref | ref | ref | ref |
| E-smoker | 1.411<br>(1.157 – 1.721)* | 1.368<br>(1.114 – 1.681)* | 1.538<br>(1.201 – 1.970)* | 1.468<br>(1.134 – 1.900)* | 0.968<br>(0.728 – 1.288) | 1.026<br>(0.765 – 1.376) |
| Dual Smoker | 1.506<br>(1.142 – 1.985)* | 1.409<br>(1.043 – 1.903)* | 1.967<br>(1.407 – 2.751)* | 1.534<br>(1.060 – 2.220)* | 1.151<br>(0.768 – 1.724) | 1.141<br>(0.738 – 1.764) |

<sup>a</sup>Adjusted for age, gender, duration of smoking, area, and financial aid
\*Significant at 0.05

## Discussion

This study provides new insights into the metabolic health profile of young Indonesian smokers, drawing on nationally representative data to examine patterns of overweight/obesity, central obesity, and hypertension across different smoking behaviors. To our knowledge, this is among the first national-level analyses to assess obesity-and blood pressure–related outcomes specifically among young users of conventional cigarettes, e-cigarettes, and dual products in Indonesia. The findings indicate that e-smokers and dual smokers consistently exhibit a higher likelihood of overweight and central adiposity compared with exclusive conventional smokers, even after adjustment for demographic characteristics. In contrast, hypertension did not differ meaningfully across smoking categories, suggesting that cardiometabolic impacts associated with smoking type may manifest more prominently in measures of adiposity than in blood pressure at this age range.

Based on our findings, subjects who reported exclusive e-cigarette use demonstrated a higher mean body mass index (BMI) compared to those who smoked conventional cigarettes or engaged in dual use. This pattern suggests that behavioral or physiological differences may exist between types of smoking exposure, particularly among young populations. A similar trend was reported by Jacobs (2018) who observed higher BMI values among e-cigarette users, although their study also noted comparable BMI levels among conventional cigarette smokers.^29^ These findings challenge the traditional perception that smoking is uniformly associated with lower body weight and instead indicate that e-cigarette use may be linked to distinct weight-related outcomes.

One possible explanation for this observation is that e-cigarette users may exhibit compensatory behaviors that differ from those of conventional smokers. For instance, the absence of combustion-related appetite suppression, combined with the perception of e-cigarettes as a “safer” alternative, may encourage increased snacking, higher caloric intake, or more sedentary lifestyles.^30^ Additionally, e-cigarette use has been associated with disordered eating behaviors among adolescents, as reported in previous studies, suggesting that psychological factors such as body image concerns, stress, or emotional regulation may also contribute to BMI among users.^31^ These adiposity-related differences appear to emerge earlier than measurable changes in blood pressure, particularly in young adults and adolescents, highlighting BMI as an early cardiometabolic marker in this population.^32^

The findings of this study are generally consistent with international evidence indicating that e-cigarette users often exhibit similar or even higher cardiometabolic risk profiles compared to conventional cigarette smokers.^33^ Several studies have reported higher BMI or increased prevalence of overweight and obesity among e-cigarette users,^34^ contrasting with earlier literature suggesting that traditional smoking suppresses appetite and is associated with lower body weight. This shift may reflect changes in nicotine delivery patterns, user behavior, and social norms surrounding e-cigarette use, particularly among adolescents.

Importantly, evidence from Southeast Asia, including Indonesia, remains limited. Most existing studies on smoking behavior and cardiometabolic risk originate from Western populations, where sociocultural contexts, dietary patterns, and regulatory environments differ substantially.^35,36^ By examining both conventional and electronic cigarette use among adolescents and young adults, this study contributes valuable regional data and helps fill an important gap in the Indonesian literature, particularly regarding early non-communicable disease (NCD) risk factors.

Several mechanisms may underlie the observed associations between smoking type, BMI, and hypertension. From a biological perspective, nicotine influences metabolic processes through activation of the sympathetic nervous system, leading to alterations in energy expenditure, insulin sensitivity, and lipid metabolism.^37^ Chronic nicotine exposure has also been linked to oxidative stress and endothelial dysfunction, which may contribute to long-term cardiometabolic risk.^38^ Although acute nicotine exposure can transiently increase metabolic rate, prolonged exposure may have paradoxical effects on weight regulation.

Behavioral mechanisms are also likely to play a significant role. E-cigarettes are often perceived as less harmful, potentially leading to more frequent use, higher cumulative nicotine exposure, and reduced motivation to maintain healthy lifestyle behaviors. Dual users, in particular, may experience greater overall nicotine intake, which could amplify both metabolic and cardiovascular effects. Additionally, sociodemographic factors such as urban residence, disposable income, access to e-cigarettes, and lifestyle characteristics may further shape these risk patterns.^39,40^

Although other unmeasured factors may influence these findings, the results highlight smoking behavior, both conventional and electronic, as an important early determinant of cardiometabolic health in adolescents. Cigarette use is associated with the early development of non-communicable disease risk factors, including overweight/obesity, hypertension, and eating disorders. Therefore, comprehensive public health policies are needed to address the rising prevalence of e-cigarette use among adolescents, strengthen prevention strategies, and ultimately reduce the long-term burden of smoking-related diseases in this vulnerable population.^41,42^

This study has several notable strengths. First, it utilizes data from the National Health Survey, which provides a large, nationally representative sample of young individuals across diverse geographic, socioeconomic, and demographic backgrounds. The use of such a comprehensive dataset enhances the external validity of the findings and enables a robust examination of how different smoking behaviors are associated with key metabolic health indicators at the population level. Second, the study focuses specifically on young smokers, a group in whom cardiometabolic changes may begin to emerge but are often underexplored in national surveys. By evaluating overweight/obesity, central obesity, and hypertension using standardized clinical cut-offs, the analysis provides insight into early metabolic risk profiles associated with conventional cigarette use, e-cigarette use, and dual use during a critical developmental period.

However, several limitations should be acknowledged. The cross-sectional design of the study restricts the ability to establish causal relationships between smoking behavior and health outcomes such as BMI, waist circumference, or blood pressure. Additionally, smoking status was based on self-report, introducing the possibility of reporting bias, as some respondents could misrepresent their smoking habits. Although the models adjusted for several important covariates, some potentially influential factors were not analyzed, including dietary intake, physical activity, sleep patterns, and psychosocial stress, all of which may contribute to adiposity and blood pressure differences. The survey also lacks detailed information on nicotine concentration, device type, or vaping frequency, which limits the ability to differentiate risk profiles within categories of e-cigarette use. Nonetheless, these limitations do not diminish the relevance of the findings, which offer nationally representative evidence on early metabolic differences among young smokers and provide an important foundation for future research and targeted prevention efforts in Indonesia.

## Conclusion

This national analysis highlights important differences in metabolic health profiles among young Indonesian smokers. While hypertension prevalence did not vary substantially across smoking categories, both e-cigarette users and dual smokers demonstrated higher odds of overweight/obesity and central obesity compared with conventional smokers, even after accounting for demographic factors. These findings suggest that alternative smoking products may be associated with early signs of metabolic risk in this age group. Considering the rapid rise of e-cigarette and dual use among Indonesian youth, these results underscore the need for early prevention strategies, integrated risk screening, and public health messaging that addresses not only the cardiovascular dangers of smoking but also its potential contribution to emerging obesity-related conditions. Continued monitoring and longitudinal research are essential to better understand how these risk patterns evolve into adulthood.

## Data Availability

The data are not publicly available due to administrative restrictions but are available from the Ministry of Health of the Republic of Indonesia upon reasonable request and official approval.

